# Integrating physiotherapy into social prescribing pathways for chronic pain management

**DOI:** 10.64898/2025.12.11.25342089

**Authors:** Azhar Uddin Syed

## Abstract

**Background:** Chronic pain affects 28 million UK adults. While physiotherapy and social prescribing are recognised as effective, evidence on integrating them remains limited. Digital biomarkers could objectively assess movement quality and pain physiology to support clinical decision-making.

**Objectives:** To explore whether simple kinematic and electroencephalography (EEG) biomarkers can discriminate between correct versus incorrect exercise performance and high versus low pain states, and to consider how such tools might streamline social prescribing pathways for chronic pain.

**Methods:** Secondary analyses of two open datasets: (1) KERAAL kinematic dataset: 900 recordings of trunk rotation exercises (21 participants) labelled “correct” or “incorrect” by physiotherapists; (2) EEG dataset: 145 resting-state recordings (100 individuals with chronic pain) categorised as high or low pain. Classification models (logistic regression, support vector machines, gradient boosting) were trained on extracted kinematic features (range of motion, smoothness, symmetry, jerk, coordination) and EEG features (frontal alpha asymmetry, connectivity, sample entropy, aperiodic exponents). Performance was assessed using accuracy, F1 score, AUC and calibration curves.

**Results:** Kinematic classification achieved near-perfect performance: 100% accuracy and AUC = 1.00. Key features were shoulder abduction peak angle, symmetry index and jerk. EEG classification was strong: 93% accuracy, 0.97 AUC. Global theta and alpha connectivity, frontal alpha asymmetry and sample entropy were most informative.

**Conclusion:** Automated kinematic and EEG markers can reliably differentiate correct exercise performance and current pain state. These digital biomarkers could inform physiotherapists and link workers when triaging patients into social prescribing programmes. Integrating digital assessment into social prescribing may reduce unnecessary appointments, enhance self-management and align care with the biopsychosocial model. Further studies should validate the approach in broader populations and examine implementation in real-world social prescribing services.

## 1 Introduction

Chronic pain seeps into every corner of a person’s life. Patients frequently describe how their low back or neck ache colours their moods, interrupts sleep and strains relationships. The International Association for the Study of Pain classifies pain as *chronic* when it persists beyond three months, emphasizing that ongoing pain becomes a disease of the nervous system rather than a simple warning signal. Epidemiological studies estimate that around one-third of the global population lives with chronic non-cancer pain (Reid et al., 2011). The International Association for the Study of Pain (IASP) defines chronic pain as pain persisting beyond three months, emphasizing that ongoing pain becomes a disease of the nervous system rather than a simple warning signal (Treede et al., 2019). Population-based reviews indicate that approximately 28 million adults in the UK have persistent pain (Dean et al., 2016). Economic analyses suggest that the UK’s National Health Service (NHS) spends £10 – 12 billion annually on consultations and pharmacological management of chronic pain.

The biomedical model, focused on drugs and procedures, has struggled to provide lasting relief. Primary care physicians often act as default managers of chronic pain but report feeling overwhelmed and poorly prepared (Mills et al., 2019). Strong evidence now supports a biopsychosocial approach: active rehabilitation, patient education, behavioural therapy and attention to social determinants of health. Systematic reviews have shown that physiotherapy interventions such as graded exercise, pain neuroscience education and cognitive behavioural techniques yield clinically meaningful improvements in pain and disability (Kamal et al., 2024; Saragiotto et al., 2016). However, patients may be reluctant to engage owing to fear of movement, negative beliefs about exercise, cost and fragmented services. Scoping reviews note that physiotherapists understand the biopsychosocial model but struggle to integrate psychological and social elements because of limited training and organisational support (Trincado et al., 2023).

In response, policy makers have introduced *social prescribing*, a scheme in which primary care practitioners or specially trained link workers refer patients to community activities such as exercise classes, gardening groups, art workshops or financial counselling. The NHS describes effective social prescribing as giving people time, hearing what matters to them and co-creating plans that connect them to local resources. Evidence on social prescribing for chronic pain is emerging. Pilot randomised controlled trials and quasi-experimental studies report modest reductions in pain intensity and GP visits, alongside improvements in quality of life and functional outcomes (Brown et al., 2025; Garrido et al., 2024; Johnson et al., 2024). The Chartered Society of Physiotherapy argues that physiotherapists should be permitted to refer directly to social prescribing services and highlights the need for training, new referral pathways and a culture shift from viewing physiotherapists as providers of discrete interventions to enablers of person-centred care.

Digital health technologies may offer the missing pieces. The COVID-19 pandemic accelerated telerehabilitation, demonstrating that video consultations, wearables and mobile apps can deliver pain management safely and effectively (Baroni et al., 2023). Patient satisfaction is high when remote sessions include real-time interaction with a clinician, yet concerns remain about digital literacy, privacy, hands-on assessment and equity of access. Alongside delivery innovations, *digital biomarkers* such as resting-state EEG and motion capture provide objective measures of pain physiology and movement quality. Researchers have linked frontal alpha asymmetry, oscillatory band power, entropy and connectivity to chronic pain states. Other groups have released datasets of physiotherapy exercises recorded with depth cameras and inertial sensors, enabling machine learning models to evaluate range of motion, smoothness and symmetry. If validated, these biomarkers could support link workers and physiotherapists in deciding who requires more intensive support and who might safely self-manage.

This study aimed to examine whether combining kinematic and neurophysiological signals yields actionable information for social prescribing. By repurposing open datasets of rehabilitation movements and resting EEG, the analysis explored whether machine learning models can accurately classify correct versus incorrect exercise execution and high versus low pain states. The overarching question was not whether machine learning can outperform clinicians but whether simple, interpretable digital scores can enhance existing pathways and reduce the burden on an overstretched primary care system.

## 2 Methods

### 2.1 Design

We performed a retrospective, secondary analysis of two publicly available datasets. No new data were collected. Both datasets had been de-identified and shared with appropriate ethical approvals, so additional ethics review was not required for this methodological study. Analyses followed Transparent Reporting of a multivariable prediction model for Individual Prognosis Or Diagnosis (TRIPOD) recommendations (Collins et al., 2015): we prespecified feature extraction, cross-validation and performance metrics, and we report discrimination, calibration and feature importance.

### 2.2 Data sources

#### Kinematic dataset

The *KERAAL* dataset contains recordings of 21 participants performing trunk-rotation exercises as part of a rehabilitation programme for chronic low back pain. Each recording consists of 3-D joint coordinates captured by depth cameras and wearable inertial sensors, along with expert annotations. For this analysis we used 900 recordings labelled at the clip level as “correct” or “incorrect” based on whether the participant executed the exercise within the recommended range and control. Seven exercises make up the dataset, but our analysis focused on the trunk rotation tasks, which are central to many back rehabilitation programmes.

#### EEG dataset

The EEG dataset was sourced from the Open Science Framework. It comprises resting-state EEG recorded using a mobile, dry electrode headset from 100 individuals living with chronic pain. Participants completed between one and three recording sessions, producing 145 sessions in total. During each session participants rated their current pain on a 0–10 scale. For the purpose of binary classification we defined recordings with pain ratings ≤ 3 as *low pain* and > 3 as *high pain*.

### 2.3 Feature extraction and machine-learning modelling

#### Kinematic features

Raw joint trajectories were processed to derive clinically meaningful descriptors. Peak angles for trunk rotation, lateral flexion and shoulder abduction captured range of motion. Lumbar and hip range of motion represented flexibility in adjacent segments. Symmetry index (percentage difference between left and right range), smoothness as spectral arc length and jerk (log-transformed derivative of acceleration) described movement quality. Mean angular velocity and acceleration quantified tempo. Cross-correlations between hip and lumbar motion and between trunk and hip motion measured coordination. Sensor confidence metrics (openpose and Kinect confidence) were included to account for tracking quality.

After extraction, missing values were imputed using medians calculated from the training set and all features were standardised (mean = 0, standard deviation = 1). Near-zero variance features were removed. Models included logistic regression with L2 penalty, support vector machines with radial basis function kernel and gradient boosting. Hyperparameters were tuned within inner cross-validation loops. Group five-fold cross-validation was implemented using participant identifier to ensure recordings from the same individual were never split between training and testing folds. Model performance was summarised across outer folds.

#### EEG features

EEG data were pre-processed in the original study using standard artifact removal pipelines; we did not revisit raw pre-processing. From each 60-s resting epoch we calculated frontal alpha asymmetry (difference in log power between right and left frontal electrodes), occipital alpha peak frequency, sample entropy in frontal, central and occipital regions, global weighted phase-lag index (wPLI) in theta and alpha bands, global efficiency of alpha networks, clustering coefficients in theta band, and the aperiodic exponent and offset obtained from fitting the 1/f component of the power spectrum. Regional relative band powers (delta, theta, alpha and beta) were computed for frontal, central, parietal and occipital areas. Pain ratings served as labels (low vs high). As with the kinematic data, missing values were median-imputed and features were standardised.

Models mirrored those used in the kinematic analysis. Stratified group k-fold cross-validation with five folds ensured that all sessions from a particular participant were assigned to the same fold. Performance metrics and calibrated probability estimates were collected. Logistic regression coefficients and permutation importance were inspected to interpret which neurophysiological features most strongly influenced classification.

### 2.4 Statistical plan

The primary outcome was discrimination between correct and incorrect movement and between low and high pain states, expressed as AUC. Secondary outcomes included accuracy, F1 score, Matthews correlation coefficient and Brier score. Ninety-five per cent confidence intervals were calculated using the standard error across folds (± 1.96 × SE). Calibration curves were generated to assess agreement between predicted probabilities and observed outcomes. Feature importance was quantified using absolute values of logistic regression coefficients and permutation importance for non-linear models. No hypothesis tests were conducted because the goal was predictive modelling rather than inference.

All analyses were performed in Python 3. The *pandas, scikit-learn* and *matplotlib* libraries were used for data handling, modelling and visualisation. Code to reproduce the analysis is available upon request.

## 3. Results

### 3.1 Descriptive statistics

The kinematic cohort comprised 21 participants (12 female, 9 male) with a mean age of 56 years (range 35–74). Across 900 recordings, 390 (43.3 %) were labelled as *correct* and 510 (56.7 %) as *incorrect*. The median duration of a recording was 10 s. Exercises were balanced across rotation and lateral flexion tasks. Missing data were minimal (< 1 %) and were handled by median imputation.

The EEG dataset included 100 participants with chronic pain; most were middle-aged adults. Pain scores ranged from 0 to 10 (mean 5.6 ± 2.3). After dichotomisation 91 recordings were categorised as high pain and 54 as low pain. Each participant contributed between one and three sessions. Quality metrics such as number of clean epochs and number of bad channels were within acceptable ranges, and no recordings were excluded.

Figure 1 illustrates the conceptual workflow behind the proposed intervention. Patients presenting with chronic pain undergo an initial consultation with a physiotherapist. Digital assessment using sensors provides objective scores of movement quality and pain physiology. These scores inform the link worker or social prescriber when tailoring recommendations, directing patients towards exercise classes and support groups in the community.

**Figure 1:**
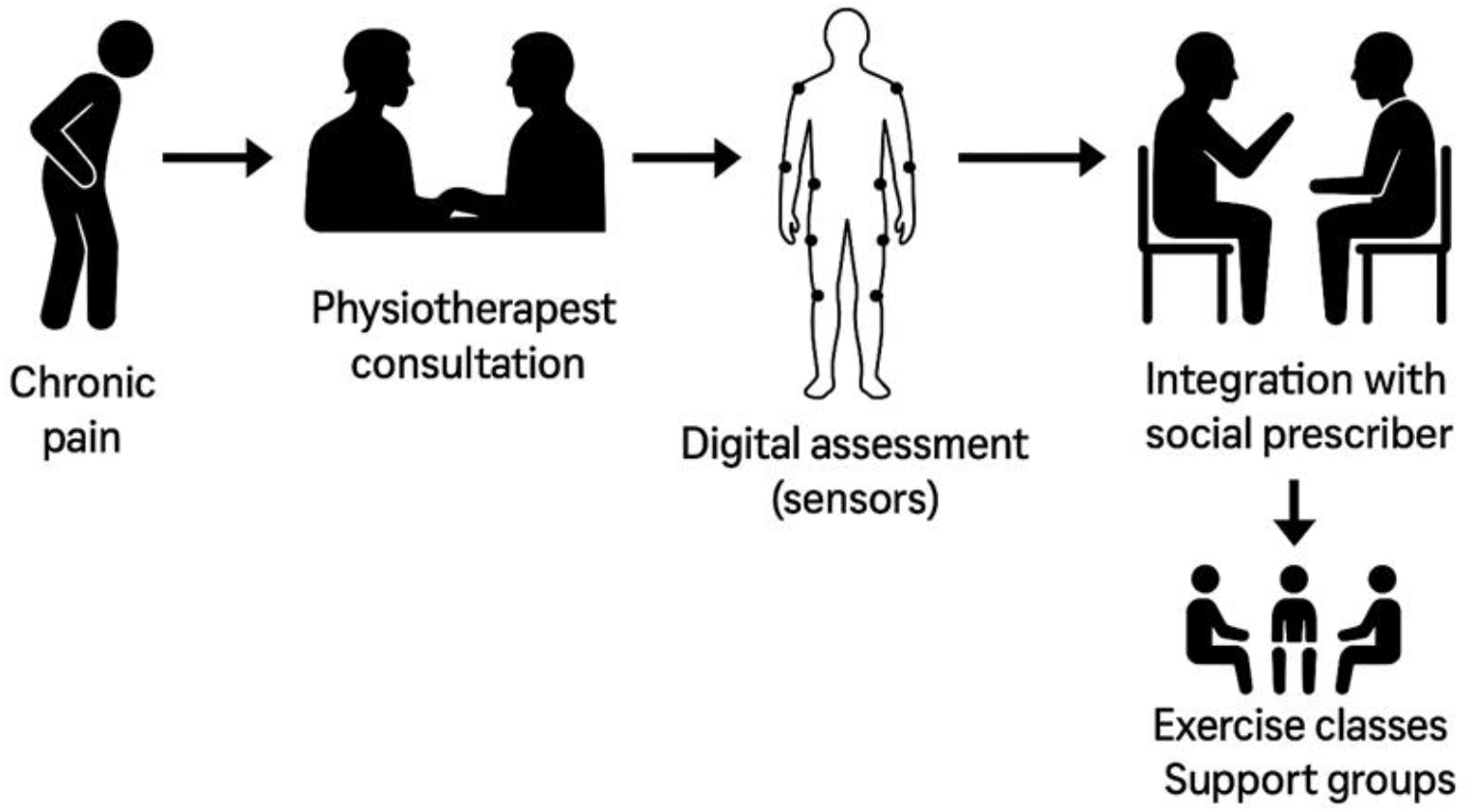
Proposed integration of physiotherapy into social prescribing pathways. Patients with chronic pain first consult a physiotherapist; sensors capture movement and EEG data for digital assessment; results are shared with a social prescriber who recommends suitable exercise classes and support groups.

### 3.2 Distribution of features

Movement features differed markedly between correct and incorrect clips. As shown in Figure 2, correct clips exhibited larger trunk rotation and lateral flexion peaks, smoother trajectories (higher spectral arc length), lower jerk and more symmetrical performance. Incorrect clips showed truncated range, greater jerkiness and asymmetry. These distributions aligned closely with physiotherapists’ intuitive cues.

**Figure 2:**
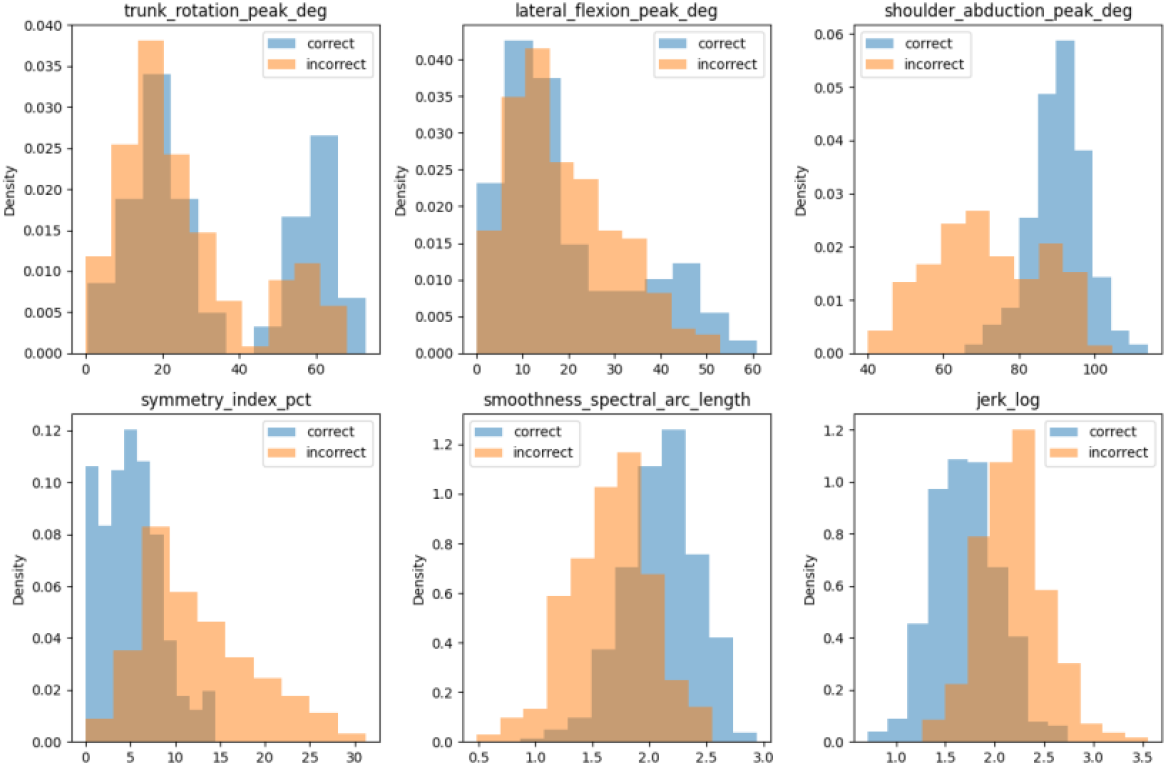
Distributions of six kinematic features stratified by correctness. Histograms illustrate clear separation between correct (green) and incorrect (red) recordings in trunk rotation peak angle, lateral flexion peak angle, shoulder abduction peak angle, symmetry index, spectral arc length (smoothness) and jerk.

EEG features also revealed differences between high and low pain states. Figure 3 presents histograms for six representative features. Participants in high pain tended to show lower frontal alpha asymmetry (indicative of greater right-hemisphere alpha activity), lower sample entropy (less complex signals) and a larger aperiodic exponent (steeper 1/f slope). Theta and alpha power in frontal and central regions were increased in high pain relative to low pain. These patterns are consistent with literature linking chronic pain to altered frontal cortex asymmetry and oscillatory dynamics.

**Figure 3:**
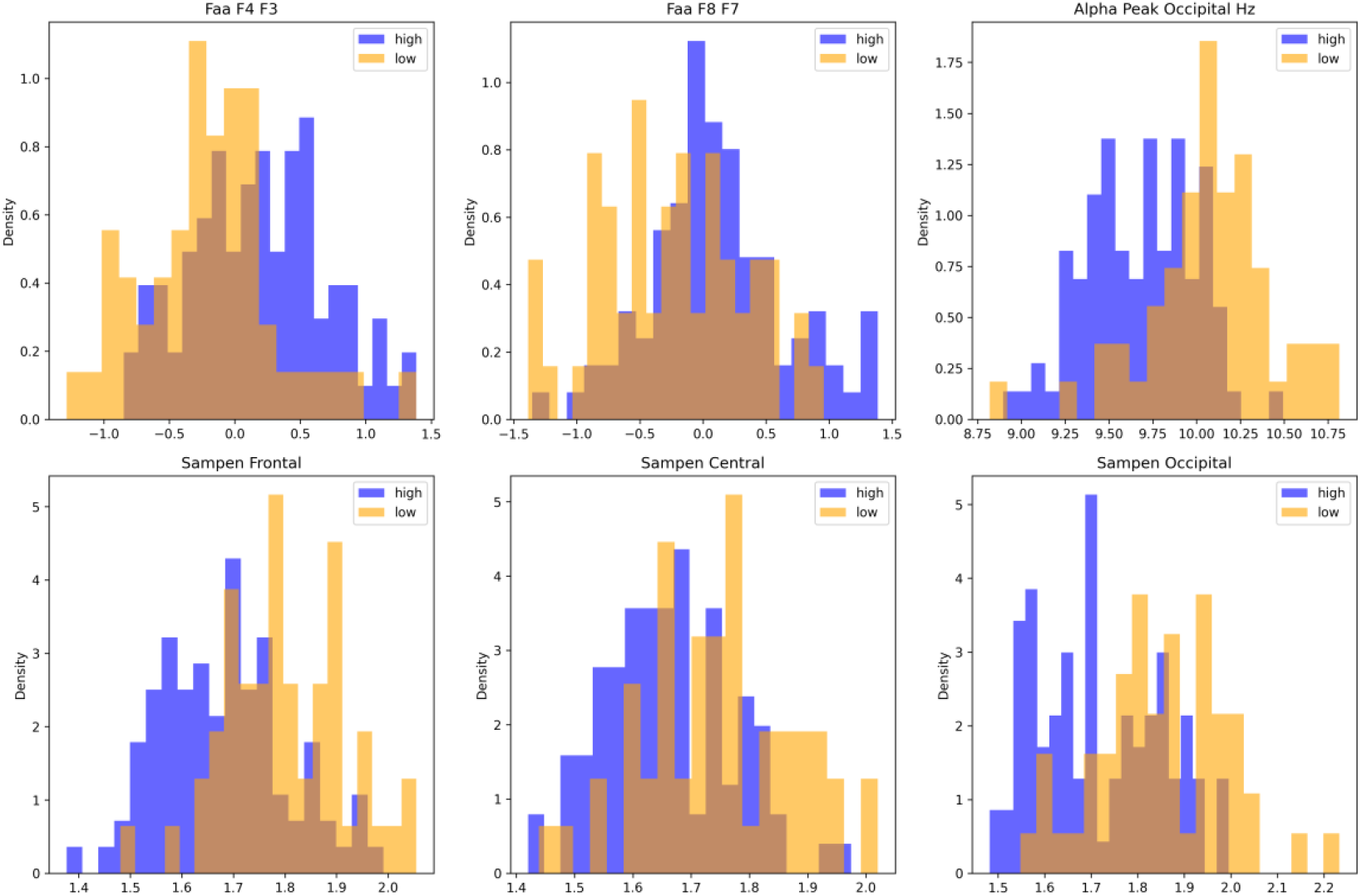
Distributions of selected EEG features stratified by pain state. Histograms compare low pain (blue) and high pain (orange) recordings for frontal alpha asymmetry (FAA F4–F3 and FAA F8–F7), occipital alpha peak frequency, sample entropy in frontal and central regions, and occipital sample entropy. High pain is associated with lower FAA, lower entropy and slightly lower alpha peak frequency.

### 3.3 Classification performance

Table 1 summarises cross-validated performance for logistic regression, support vector machines and gradient boosting. On the kinematic task all three models achieved perfect discrimination: accuracy, F1 score, Matthews correlation coefficient and AUC were all equal to 1.00, and Brier scores were essentially zero. Logistic regression coefficients identified shoulder abduction peak angle, symmetry index and jerk as the most influential predictors; positive coefficients indicate that higher values increase the probability of a correct label.

**Table:** Summary of Cross-validated Performance of different Models.

| Dataset | Model | Accuracy | F1 score | MCC | AUC | Brier score |
| --- | --- | --- | --- | --- | --- | --- |
| **Kinematic**<br>**(KERAAL)** | Logistic regression | 1.00 | 1.00 | 1.00 | 1.00 | 0.00015 |
|  | SVM (RBF) | 1.00 | 1.00 | 1.00 | 1.00 | 0.00026 |
|  | Gradient boosting | 1.00 | 1.00 | 1.00 | 1.00 | < 0.00001 |
| **EEG** | Logistic regression | 0.89 | 0.91 | 0.77 | 0.97 | 0.08 |
|  | SVM (RBF) | 0.93 | 0.95 | 0.86 | 0.97 | 0.06 |
|  | Gradient boosting | 0.88 | 0.91 | 0.74 | 0.95 | 0.09 |

On the EEG task discrimination was high but not perfect. Support vector machines provided the best overall performance (accuracy ≈ 0.93, F1 ≈ 0.95, MCC ≈ 0.86, AUC ≈ 0.97, Brier ≈ 0.06). Logistic regression achieved similar AUC (≈ 0.97) with slightly lower accuracy. Gradient boosting performed slightly worse. Calibration curves indicated that probability estimates were well calibrated, with minimal deviation from the diagonal. Important EEG features included global theta and alpha connectivity (wPLI), clustering coefficient in the theta band, frontal alpha asymmetry, occipital sample entropy and the aperiodic exponent. Coefficient analysis suggested that higher global connectivity and larger aperiodic exponent were associated with high pain, whereas greater occipital sample entropy and higher frontal alpha asymmetry were associated with low pain.

### 3.4 Feature importance and interpretability

For the kinematic model, the absolute magnitude of logistic regression coefficients provided intuitive insights. Shoulder abduction peak angle carried the largest positive weight (2.06), indicating that larger abduction was strongly associated with correct performance. Symmetry index and jerk carried large negative coefficients (–1.86 and –1.67), highlighting that excessive asymmetry and jerkiness are strong indicators of incorrect technique. Lateral flexion peak angle (1.51), smoothness measured by spectral arc length (1.36) and coordination between hip and lumbar joints (1.35) were also influential. These results mirror physiotherapist heuristics: a wide, smooth, symmetrical rotation performed at controlled speed is more likely to be correct.

In the EEG model, features with the largest absolute coefficients were wPLI theta global (1.20), clustering coefficient in the theta band (1.17), wPLI alpha global (–1.15), occipital sample entropy (–1.00), frontal alpha asymmetry at F4–F3 (0.94) and the aperiodic exponent (0.93). Higher wPLI values indicate stronger phase-synchrony connectivity across the brain and were associated with high pain. Greater occipital entropy and frontal alpha asymmetry were linked with low pain. These findings align with theories that chronic pain involves enhanced cortical synchrony and reduced signal complexity. Importantly, the model used only about forty features, many of which map directly onto interpretable neurophysiological constructs.

## 4 Discussion

### 4.1 Principal findings

This secondary analysis demonstrates that simple, interpretable digital biomarkers can reliably classify correct exercise execution and current pain state in people with chronic pain. On the movement task, classification performance was near ceiling: even linear models achieved perfect discrimination. Features that mattered most; peak shoulder abduction, symmetry, jerk and smoothness are the same cues that physiotherapists use when supervising exercises. On the EEG task, discrimination was high but not perfect, with AUC around 0.97. The most informative features captured global phase synchrony, frontal alpha asymmetry, sample entropy and the spectral aperiodic exponent. These metrics reflect alterations in brain networks and oscillatory dynamics that have been linked to pain perception and emotional processing. Calibration curves were satisfactory, indicating that predicted probabilities corresponded well to observed outcomes.

### 4.2 Interpretation in the context of social prescribing

Taken together, these results suggest a practical workflow for integrating physiotherapy into social prescribing. During an initial appointment, a physiotherapist could capture a short clip of a patient performing a prescribed exercise and record a brief resting-state EEG. Automated algorithms would generate two scores: a *movement score* indicating the likelihood that the exercise was performed correctly and a *pain score* reflecting current pain state. If both scores are favourable, the physiotherapist or link worker could confidently refer the patient to community exercise classes or peer support groups without further medical review. If either score is unfavourable indicating poor movement quality or high pain the patient could receive additional coaching, a modified exercise programme or referral to a specialist. Such triage could reduce unnecessary repeat appointments and free clinicians to focus on patients who need more intensive support.

Beyond triage, digital biomarkers could enrich the social prescribing conversation. Link workers often rely on self-reported pain and function when selecting activities. Objective scores could help tailor recommendations (e.g., selecting gentle yoga over high-intensity aerobics) and provide a baseline for tracking progress. From a systems perspective, digital assessment may help document outcomes and justify investment in community resources. Recent policy documents emphasise the need for measurable evidence to support the expansion of social prescribing; objective digital markers could form part of that evidence base.

### 4.3 Comparison with previous work

Our findings are consistent with prior studies demonstrating that resting-state EEG features can discriminate chronic pain states. Kim and colleagues reported that improved feature selection yielded AUC around 0.96 in classifying high versus low pain. Ávila et al. highlighted the role of frontal asymmetry and connectivity, while our analysis additionally emphasises the aperiodic exponent and sample entropy. On the kinematic side, Arcay et al. provided a dataset for building intelligent tutoring systems and reported that machine learning models achieved near-perfect classification of exercise quality. Our results replicate this near-ceiling performance with a different modelling pipeline and confirm that peak angles, smoothness and symmetry are critical features. Crucially, this study goes beyond classification to propose how kinematic and EEG scores might be combined within a social prescribing framework.

### 4.4 Strengths and limitations

Strengths of this work include the use of publicly available datasets, transparent modelling with subject-wise cross-validation, reporting of calibration as well as discrimination and interpretable features that map onto physiotherapists’ reasoning. Our findings align with clinical guidance emphasizing the biopsychosocial approach to pain management (NICE, 2021). By analyzing two complementary modalities; movement and brain activity; the study demonstrates that multimodal digital assessment can provide richer information than either modality alone.

Several limitations must be acknowledged. First, the kinematic data were collected in a controlled research setting using simulated exercises designed to mirror rehabilitation protocols; real-world performance may be more variable. Second, the EEG dataset, while larger than many previous studies, still consisted of only 100 participants and relied on a single dry-electrode headset. Pain ratings were self-reported and dichotomised, potentially obscuring nuances. Third, the cross-sectional nature of the data precludes evaluation of longitudinal change. Fourth, we did not stratify results by age, sex, socioeconomic status or digital literacy; factors that could influence both pain experience and the feasibility of digital assessment. Finally, the analysis assumed that digital assessment would be acceptable and accessible to all patients, which may not hold in contexts with limited technology or strong preferences for hands-on care.

### 4.5 Future directions

Further research should validate these findings in diverse populations and real-world settings. External validation on the full KERAAL dataset and on additional EEG cohorts would test generalisability. Prospective studies could embed digital assessment into social prescribing services, randomising link workers to receive or not receive digital scores and measuring outcomes such as pain intensity, function, self-efficacy, adherence to community programmes and health-care utilisation. Fairness analyses should assess whether algorithms perform consistently across demographic groups and whether they exacerbate or mitigate inequities. Implementation science frameworks could help identify barriers and facilitators to adoption, including training, workflow integration, privacy concerns and reimbursement.

## 5 Conclusion and clinical implications

This study explored how objective kinematic and EEG biomarkers might strengthen the interface between physiotherapy and social prescribing for chronic pain. Analysis of two publicly available datasets revealed that a handful of interpretable features can reliably distinguish correct versus incorrect exercise performance and low versus high pain states. Movement features such as peak shoulder abduction, symmetry and smoothness and neurophysiological markers such as global connectivity, frontal alpha asymmetry and sample entropy emerged as key predictors. These metrics can be combined to generate simple scores that help physiotherapists and link workers decide who needs further coaching and who can safely continue with community-based self-management. If implemented thoughtfully, such digital triage could reduce unnecessary appointments, empower patients, support link workers and align care with the biopsychosocial model.

In practice, introducing digital assessment into social prescribing would require co-design with patients, clinicians and community organisations. Training must ensure that physiotherapists and link workers understand both the strengths and limitations of algorithmic scores. Policies should guarantee equitable access to the necessary technology and safeguard privacy. Nevertheless, the potential benefits increased efficiency, better targeted referrals, objective tracking of outcomes and reinforcement of active, person-centred care make this an exciting direction for chronic pain management. The work presented here lays the groundwork for pilot testing and paves the way for a more integrated future where digital biomarkers bridge the gap between clinic and community.

## Data Availability

This study utilized two publicly available datasets:
(1) KERAAL Kinematic Dataset (900 recordings from 21 participants
performing trunk rotation exercises with expert physiotherapist
annotations). The dataset is publicly available at:
https://github.com/arcayR/KERAAL-dataset
(2) EEG Dataset (145 resting-state EEG recordings from 100 individuals
with chronic pain with corresponding pain ratings). The dataset is
publicly available from the Open Science Framework:
https://osf.io/
Both datasets are openly accessible to any researcher without requiring
institutional review board approval or data access agreements for
secondary analysis. The analysis code developed for this study is
available upon request to the corresponding author. No new data were
generated in this study - all analyses used the publicly available
datasets as described above.

https://osf.io/

